# Predictive Modeling to Uncover Parkinson’s Disease Characteristics That Delay Diagnosis

**DOI:** 10.1101/2024.03.12.24304165

**Authors:** Tom Hähnel, Tamara Raschka, Jochen Klucken, Enrico Glaab, Jean-Christophe Corvol, Björn H. Falkenburger, Holger Fröhlich

**Author notes:** <u>Corresponding author:</u> Dr. Tom Hähnel, Department of Neurology, Medical Faculty and University Hospital Carl Gustav Carus, Dresden University of Technology, Fetscherstr. 74, 01307 Dresden, Germany, Email: haehnto.accounts+. **Funding sources:** The project has been partially funded by the ERA PerMed EU-wide project DIGIPD (01KU2110) and the ParKInsonPredict project (to TH, 16DKWN1113A) funded by the Federal Ministry of Education and Research of Germany (Bundesministerium für Bildung und Forschung). The ICEBERG Study was funded by the Programme d’investissements d’avenir (ANR-10-IAIHU-06), the Paris Institute of Neurosciences – IHU (IAIHU-06), the Agence Nationale de la Recherche (ANR-11-INBS-0006), and Électricité de France (Fondation d’Entreprise EDF). LuxPARK is part of the National Centre of Excellence in Research on Parkinson’s Disease (NCER-PD), which is funded by the Luxembourg National Research Fund (FNR/NCER13/BM/11264123, INTER/ERAPerMed 20/14599012).

## Abstract

**Background:** People with Parkinson’s disease (PwPD) present with a variety of motor and non-motor symptoms, and a more biological definition of PD is poised to expand the diagnostic spectrum beyond the stereotypical “elderly male with tremor”. This heterogeneity can potentially pose a challenge for an accurate and early diagnosis.

**Objectives:** To determine whether demographic or clinical characteristics systematically affect the time till diagnosis, by modeling large-scale longitudinal data.

**Methods:** Using longitudinal data from three large PD cohorts and a latent time joint mixed-effects model (LTJMM), we aligned the disease courses of individual PwPD and estimated whether individual PD diagnosis was early or late compared to the average time of PD diagnosis in each cohort. Initial clinical manifestations at the typical time of PD diagnosis were estimated using mixed-effects models.

**Results:** We included 1,124 PwPD in our analysis. Several clinical and demographic factors were associated with a later-than-average diagnosis of PD: higher age, tremor dominance, rapid progression, anxiety, autonomic symptoms, depression, fatigue, pain, sleep problems, and in general more non-motor symptoms. In contrast, postural and gait disturbance was associated with an earlier-than-average PD diagnosis. Sex, family history of PD and predominantly affected side did not impact the time of PD diagnosis.

**Conclusions:** Using statistical modeling, we were able to study initial clinical characteristics of PwPD even in the absence of directly observable clinical data at the time when PD is diagnosed typically. Our findings are consistent with a biological definition of PD that includes patients who present initially with non-motor symptoms.

## Introduction

Parkinson’s disease (PD) is the fastest-growing neurological disease and the second most common neurodegenerative disease (1). Numerous ongoing clinical trials investigate potentially disease-modifying treatments for PD aiming to stop or significantly slow neurodegeneration (2). This development outlines the increasing importance of early and accurate diagnosis in PD, as timely interventions will be required to maximize the therapeutic benefits of these emerging treatments. However, while the stereotypical person with Parkinson’s disease (PwPD) is an elderly patient with tremor (3,4), the clinical picture of motor and non-motor symptoms in early PD is much more diverse. This heterogeneity challenges early diagnosis in PD (5–7). It is therefore crucial to identify clinical features associated with delayed diagnosis in PD.

The exploration of diagnostic delay is challenging because structured clinical data preceding the formal PD diagnosis are generally not available. Prior studies have utilized the interval between self-recognition of motor symptoms and the clinical PD diagnosis as a surrogate measure of diagnostic delay. Clinical characteristics in these studies were assessed either through structured clinical assessments at study inclusion (8–11) or retrospectively, using general information obtained from electronic health records or registers (6,12). Results from these studies have yielded inconsistent findings with regard to demographic factors (8–13). Interestingly, some studies indicated that the presence of non-motor symptoms may contribute to a delay in PD diagnosis (6,10). However, a detailed investigation of which specific non-motor symptoms are responsible for this diagnostic delay remains unexplored.

Our study aims to objectively and systematically identify clinical and demographic factors associated with diagnostic delay in PD, by using longitudinal data from three large PD cohorts. To achieve this, we propose a novel, model-based methodology designed (I) to objectively estimate the diagnostic delay for individual PwPD in comparison to the average time when PD is diagnosed, and (II) to infer the initial clinical manifestations of individual PwPD from longitudinal data, even in the absence of clinical information before study inclusion. Based on these estimates, we systematically studied which demographic and clinical factors influence diagnostic delay in PD.

## Methods

### Clinical cohorts

We included PwPD from three distinct cohort studies into our analysis: (I) the Parkinson’s Progression Markers Initiative (PPMI, NCT04477785) (14), which focuses on de-novo PwPD, (II) the French ICEBERG cohort study (NCT02305147) with early-stage PwPD, and (III) the Luxembourg Parkinson’s Study (LuxPARK, NCT05266872) (15) comprising PwPD across all disease stages. Details regarding the inclusion and exclusion criteria can be found in the supplementary material. All study groups have obtained approval from their respective ethical committees and participant consent aligned with the Declaration of Helsinki.

### Motor phenotypes

PwPD were categorized as tremor dominant (TD) or postural instability and gait disturbance (PIGD) subtypes based on the ratio of the respective Unified Parkinson’s Disease Rating Scale (UPDRS) part III sub-scores (16).

### Estimating time shifts

To estimate the diagnostic delay of individual PwPD diagnoses, we calculated how much each patient’s timescale is shifted from the average PD time course. Therefore, we utilized a latent time joint mixed-effects model (LTJMM) (17). In brief, LTJMM models linear progression of multiple clinical outcomes jointly over time, thereby estimating the clinical course of a mean PwPD and how individual PwPD are shifted from this average PD time course. Thereby, LTJMM includes random effects to accommodate for individual differences in progression speed of clinical outcomes. These model-derived time shifts were used to align PwPD on a *common disease timescale* and reflect how early or late individual PwPD were diagnosed compared to an average PwPD. We included clinical scores assessing motor and non-motor symptoms as outcomes into the LTJMM: UPDRS I-IV, PIGD, Montreal Cognitive Assessment (MoCA) and Scales for Outcomes in Parkinson’s Disease-Autonomic Dysfunction (SCOPA). We accounted for age and sex as potential confounding covariates. The LTJMM was implemented via the R packages ltjmm (18) and RStan (19). Further details can be found in the supplementary material.

### Estimating initial clinical manifestations

After aligning PwPD on the common disease timescale, we predicted the clinical manifestations of individual PwPD at the time when PD would be expected to be diagnosed. Therefore, we predicted the initial values of 66 outcomes (including single questions, scores and sub-scores from questionnaires and clinical assessments, Table S1) on the common disease timescale, i.e. at the time where PD is diagnosed on average. This was achieved by using linear mixed-effects models, binary mixed-effects models and ordinal mixed-effects models depending on the scales of the outcomes. Subsequently, we calculated the correlation of the predicted initial clinical outcomes with (I) patient-reported time to diagnosis and (II) the model-derived time shifts. Depending on the scale of the outcome, Pearson, Kendall Tau-b or point-biserial correlation was used.

To enable comparability among the three cohorts, each utilizing different clinical scores, we used meta-analyses to aggregate correlations on the level of symptom domains rather than using individual scores (Table S1).

Finally, p-values and confidence intervals (CIs) were adjusted for multiple testing across all 17 symptom domains using the Benjamini-Hochberg procedure (20). Details are provided in the supplement.

### Progression subtypes

We aimed to compare whether the rate of disease progression impacts how early or late PD is diagnosed by comparing two subtypes identified in a recent publication: a fast-progressing and a slow-progressing subtype (21). Using the same approach as described in our previous publication, i.e. variational deep embedding with recurrence (VADER) (22), we assigned all PwPD to one of these two subtypes.

### Statistical analyses

For comparison of cohort characteristics, the following tests were applied: Sex was compared using Fisher’s exact test, Hoehn & Yahr using the Kolmogorov-Smirnov test and all other characteristics were compared using the Mann-Whitney U test. P-values were adjusted for multiple testing using the Benjamini-Hochberg procedure (20).

Correlations of patient-reported time since diagnosis and model-derived time shifts with age of diagnosis were calculated using Pearson correlation. Group differences regarding sex, family history, motor phenotype and predominant side were carried out using t-test, Welch’s test or Mann-Whitney U test depending on the distribution of the data. Subgroup analyses for sex and age-specific subgroups were conducted for characteristics with significant associations in the overall analysis. The age-specific subgroups were defined by a median split on age of diagnosis covering PwPD from all three cohorts.

All statistical tests were conducted as two-tailed tests with significance level 0.05 using the Python package SciPy (23).

### Code and data availability

Relevant source code will be published at https://github.com/SCAI-BIO/PD-diagnostic-delay upon acceptance of the paper. Availability of the clinical data depends on the individual study groups.

## Results

### Demographic and clinical characteristics

A total of 1,124 PwPD from three cohorts were included into our analysis. The cohorts exhibited differences in clinical presentation at baseline, largely associated with differences in disease duration between the cohorts. Specifically, the LuxPARK cohort was predominantly composed of PwPD at more advanced disease stages compared with ICEBERG and PPMI, which primarily included early stage PwPD (Table **1**).

**Table 1:**
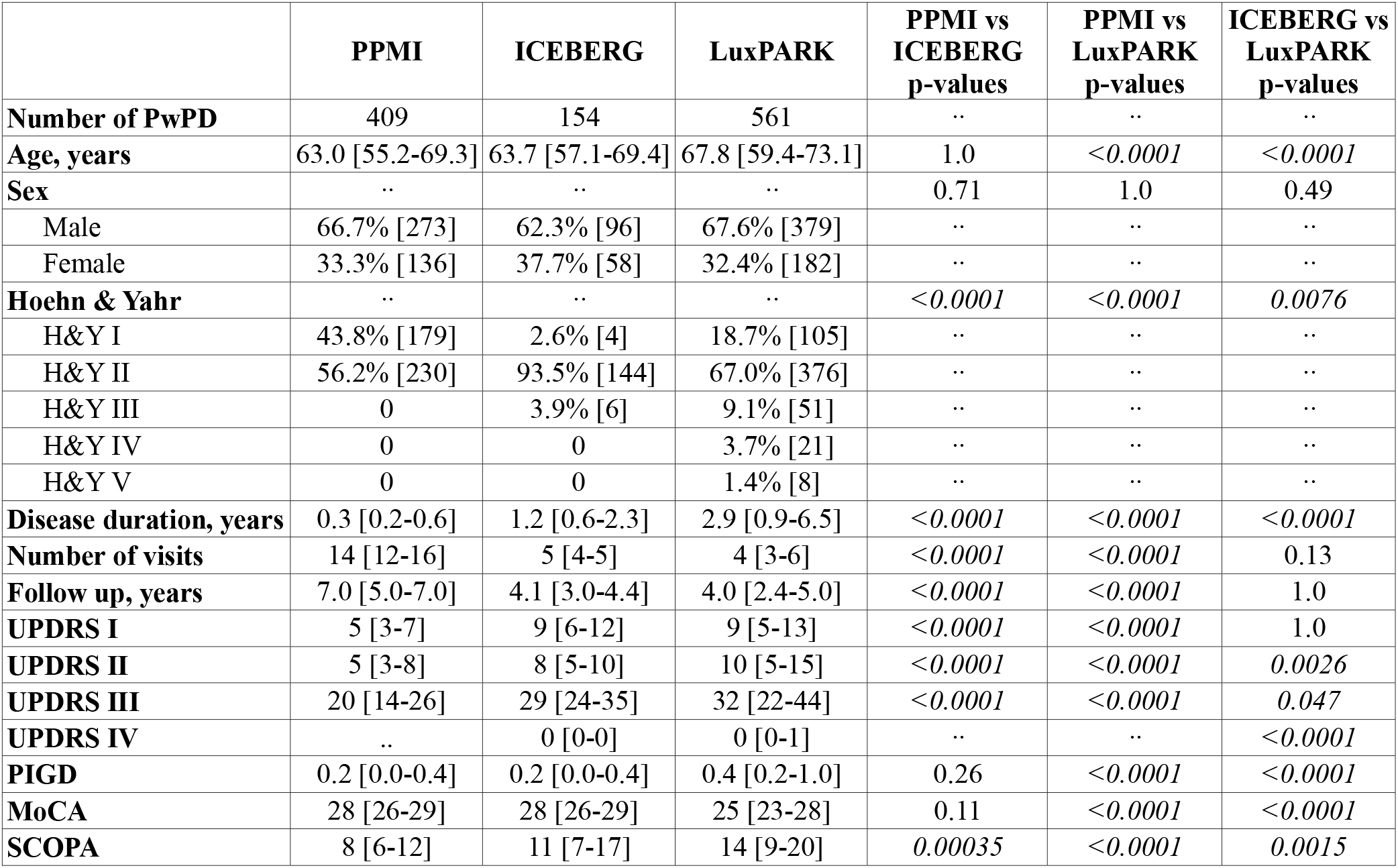
Baseline demographic and clinical characteristics of PPMI, ICEBERG and LuxPARK cohorts. Relative and absolute frequencies (sex and H&Y), respective medians and interquartile ranges (other outcomes) are shown.

|  | PPMI | ICEBERG | LuxPARK | PPMI vs<br>ICEBERG<br>p-values | PPMI vs<br>LuxPARK<br>p-values | ICEBERG vs<br>LuxPARK<br>p-values |
| --- | --- | --- | --- | --- | --- | --- |
| **Number of PwPD** | 409 | 154 | 561 | .. | .. | .. |
| **Age, years** | 63.0 [55.2-69.3] | 63.7 [57.1-69.4] | 67.8 [59.4-73.1] | 1.0 | <0.0001 | <0.0001 |
| **Sex** | .. | .. | .. | 0.71 | 1.0 | 0.49 |
| Male | 66.7% [273] | 62.3% [96] | 67.6% [379] | .. | .. | .. |
| Female | 33.3% [136] | 37.7% [58] | 32.4% [182] | .. | .. | .. |
| **Hoehn & Yahr** | .. | .. | .. | <0.0001 | <0.0001 | 0.0076 |
| H&Y I | 43.8% [179] | 2.6% [4] | 18.7% [105] | .. | .. | .. |
| H&Y II | 56.2% [230] | 93.5% [144] | 67.0% [376] | .. | .. | .. |
| H&Y III | 0 | 3.9% [6] | 9.1% [51] | .. | .. | .. |
| H&Y IV | 0 | 0 | 3.7% [21] | .. | .. | .. |
| H&Y V | 0 | 0 | 1.4% [8] | .. | .. | .. |
| **Disease duration, years** | 0.3 [0.2-0.6] | 1.2 [0.6-2.3] | 2.9 [0.9-6.5] | <0.0001 | <0.0001 | <0.0001 |
| **Number of visits** | 14 [12-16] | 5 [4-5] | 4 [3-6] | <0.0001 | <0.0001 | 0.13 |
| **Follow up, years** | 7.0 [5.0-7.0] | 4.1 [3.0-4.4] | 4.0 [2.4-5.0] | <0.0001 | <0.0001 | 1.0 |
| **UPDRS I** | 5 [3-7] | 9 [6-12] | 9 [5-13] | <0.0001 | <0.0001 | 1.0 |
| **UPDRS II** | 5 [3-8] | 8 [5-10] | 10 [5-15] | <0.0001 | <0.0001 | 0.0026 |
| **UPDRS III** | 20 [14-26] | 29 [24-35] | 32 [22-44] | <0.0001 | <0.0001 | 0.047 |
| **UPDRS IV** | .. | 0 [0-0] | 0 [0-1] | .. | .. | <0.0001 |
| **PIGD** | 0.2 [0.0-0.4] | 0.2 [0.0-0.4] | 0.4 [0.2-1.0] | 0.26 | <0.0001 | <0.0001 |
| **MoCA** | 28 [26-29] | 28 [26-29] | 25 [23-28] | 0.11 | <0.0001 | <0.0001 |
| **SCOPA** | 8 [6-12] | 11 [7-17] | 14 [9-20] | 0.00035 | <0.0001 | 0.0015 |

### Patient-reported time to diagnosis

Measuring diagnostic delay is complicated by the fact that data from the time before diagnosis is not available. In addition, there is a delay between recognition of first symptoms and diagnosis, but also a delay between diagnosis and inclusion into longitudinal cohorts with systematic assessment of symptoms. Thus, we can only directly assess (I) the time elapsed between the first motor symptom self-recognized by PwPD and PD diagnosis, and (II) the clinical characteristics of the PwPD at the first study visit. We will refer to this time span between recognition of first motor symptom and PD diagnosis as *patient-reported time to diagnosis*. Median patient-reported time to diagnosis was 0.9 years for PPMI, 1.1 years for ICEBERG and 1.0 years for LuxPARK (Fig. S1), being in line with previous publications using a similar approach (8–12,24).

Subsequently, we explored how demographic characteristics influenced patient-reported time to diagnosis, thereby finding no differences based on sex, age of diagnosis, family history, predominant PD side and motor phenotype (Table S2).

To gain deeper insights whether specific PD symptoms are associated with a longer time to diagnosis, we evaluated several clinical symptom domains at baseline visit by incorporating various questionnaires and assessments from the different cohorts (Table S1). We used only PPMI and ICEBERG visits for this analysis as they included newly diagnosed PwPD in contrast to LuxPARK including PwPD from all disease stages. Following this approach, we observed no significant associations of patient-reported time to diagnosis with baseline clinical scores (Fig. S2, Table S3).

### Model-based estimation of time shifts and initial manifestations

Subsequently, we used a model-based approach to objectively estimate diagnostic delay and initial clinical manifestations at a comparable time. Specifically, a latent-time joint mixed-effects model (LTJMM) was fitted on trajectories of motor and non-motor scores of individual PwPD in the three cohorts (Fig. **1**A). This model was originally designed to account for different disease stages of patients in longitudinal studies by aligning symptom trajectories on a common timescale. As a first step, the model calculates for each PwPD how much the clinical course differs from the average PD time course estimated from all PwPD in the cohort. The model thus provides symptom trajectories aligned on a common disease timescale, i.e. the disease time course of a typical PwPD. In addition, the model provides for each PwPD a positive or negative time shift between their trajectory and the common disease timescale. Since the individual time of diagnosis is known in all PwPD, this modeling approach allows us to determine whether patients were diagnosed earlier or later than average, providing us an estimate of the diagnostic delay for each PwPD (Fig. **1**B).

**Figure 1:**
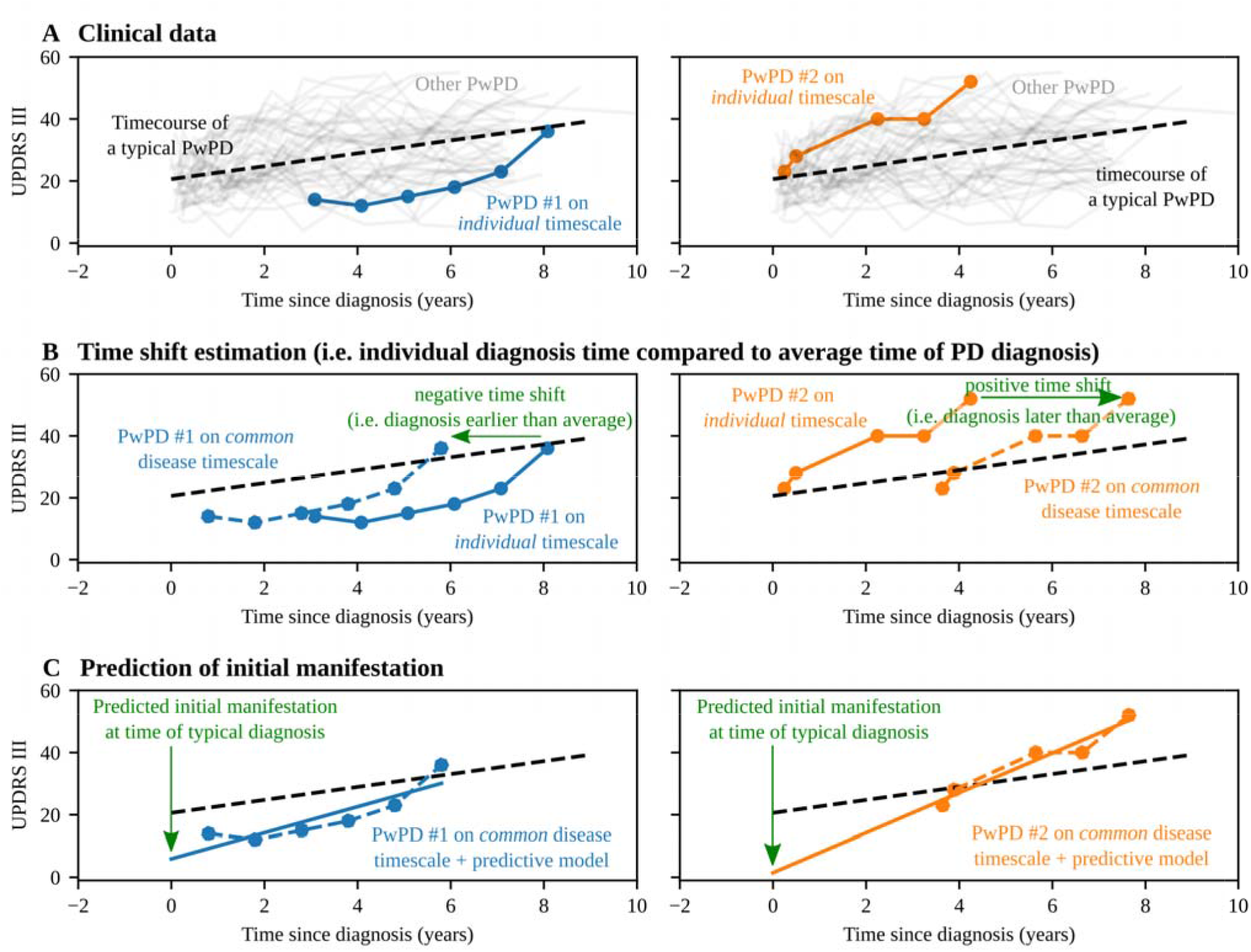
Methodology for calculating model-derived time shifts and initial manifestations of PwPD.

Subsequently, we used statistical models to estimate the initial clinical manifestations of individual PwPD, i.e., the symptoms exhibited at the time when PD would be typically diagnosed (Fig. **1**C). Validations of the LTJMM and the statistical models are presented in the supplement (Fig. S3 and S4).

As expected, the distribution of model-derived time shifts centered around zero years for each cohort with standard deviations of 1.7 (PPMI), 3.5 (ICEBERG), and 3.6 years (LuxPARK) (Fig. S5). We observed a mild correlation between patient-reported time to diagnosis and model-derived time shifts for ICEBERG (_ρ_=0.16, P=0.045) while there was no significant correlation for PPMI, LuxPARK and in the pooled analysis (Fig. S6).

### Demographic factors determining a late diagnosis

We subsequently explored which demographic attributes were associated with early PD diagnosis (negative model-derived time shift) and late PD diagnosis (positive model-derived time shift).

Neither sex, family history of PD nor predominant PD side showed an association with model-derived time shifts (Table S4), in line with our first analysis of patient-reported time to diagnosis (Table S2). Adjusting the age of diagnosis by the model-derived time shifts allowed us to effectively estimate the age when PD would have been expected to be diagnosed in individual PwPD. With this approach, we observed a strong correlation of higher predicted age at diagnosis with later-than-average PD diagnosis (reported by positive model-derived time shifts). This finding was consistent across all three cohorts (Fig. **2**, Table S4) and for the pooled analysis (_ρ_=0.56, P<0.0001). It indicates that PD symptoms could be recognized less in older patients, leading to a delayed diagnosis.

**Figure 2:**
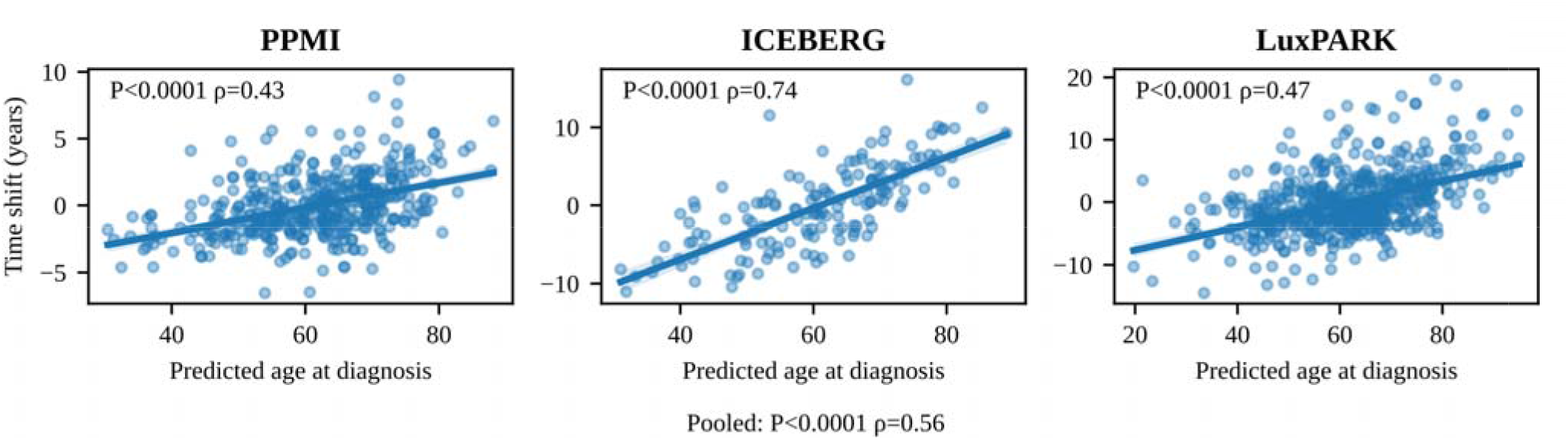
Correlation of estimated age at diagnosis with model-derived time shift. Positive model-derived time shifts indicate that PD was diagnosed later than average.

Looking at sex-specific subgroups, similar correlations of higher predicted age at diagnosis with later-than-average PD diagnosis were reproducible for both male (pooled estimate: _ρ_=0.60, P<0.0001) and female (pooled estimate: _ρ_=0.48, P=0.0015) PwPD (Table S4).

### Clinical symptoms determining a late diagnosis

To gain deeper insights into the initial clinical manifestation of PwPD associated with a later-than-average PD diagnosis, we evaluated several clinical symptom domains using information from questionnaires and assessments included in the three cohorts (Table S1). Instead of analyzing the clinical manifestations at baseline visit, we predicted the clinical manifestations at the time when PD would be expected to be diagnosed (i.e. if it was not delayed, Fig. **1**C).

Several non-motor symptoms were associated with a later-than-average diagnosis of PD (Fig. **3**, Table S5) while higher axial and PIGD symptoms were linked with an earlier-than-average PD diagnosis. These findings were consistent across all three cohorts, as detailed in the forest plots provided in the supplementary data. They indicate that PD could be suspected less in patients presenting predominantly with non-motor symptoms, which could lead to a delayed diagnosis of PD.

**Figure 3:**
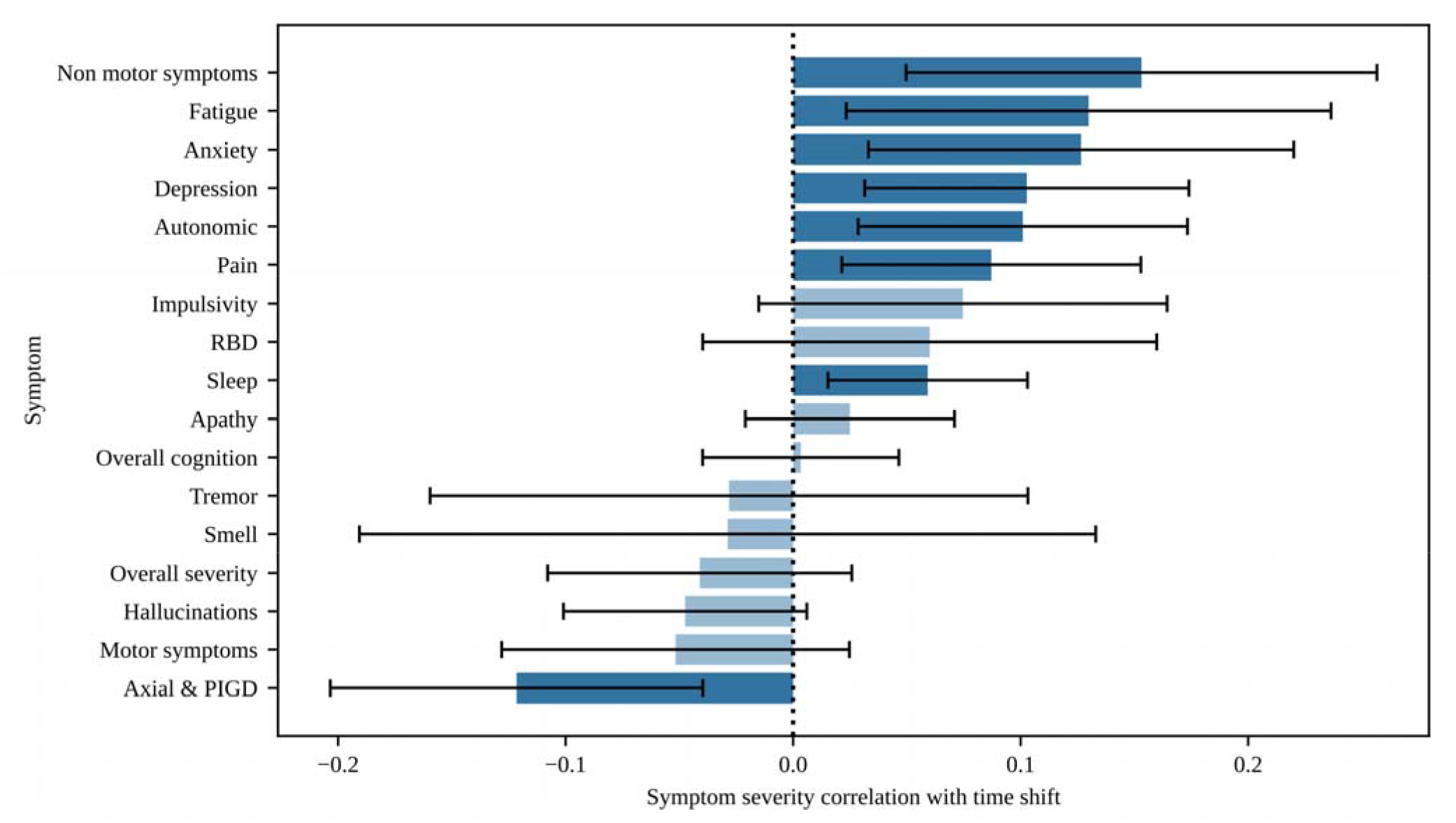
Correlations between initial clinical characteristics and model-derived time shift. Positive correlation coefficients indicate that an increased symptom severity is associated with a PD diagnosis later than average.

Exploring sex-specific variations, we noted several differences compared to the pooled analysis. For male PwPD, neither pain nor sleep disturbances influenced the time of PD diagnosis significantly. In contrast, female PwPD demonstrated an association of impulsivity with a later-than-average PD diagnosis (Table S5).

Surprisingly, PD diagnosis was later than average in PwPD with TD subtype than in PwPD with PIGD subtype. This observation held for the PPMI and LuxPARK cohorts and the pooled analysis (d=0.35, P=0.0005), but was not statistically significant for the smaller ICEBERG cohort (Fig. **4**A, Table S4). The association of later-than-average PD diagnosis in PwPD of TD subtype was consistent in both sex-specific subgroups.

**Figure 4:**
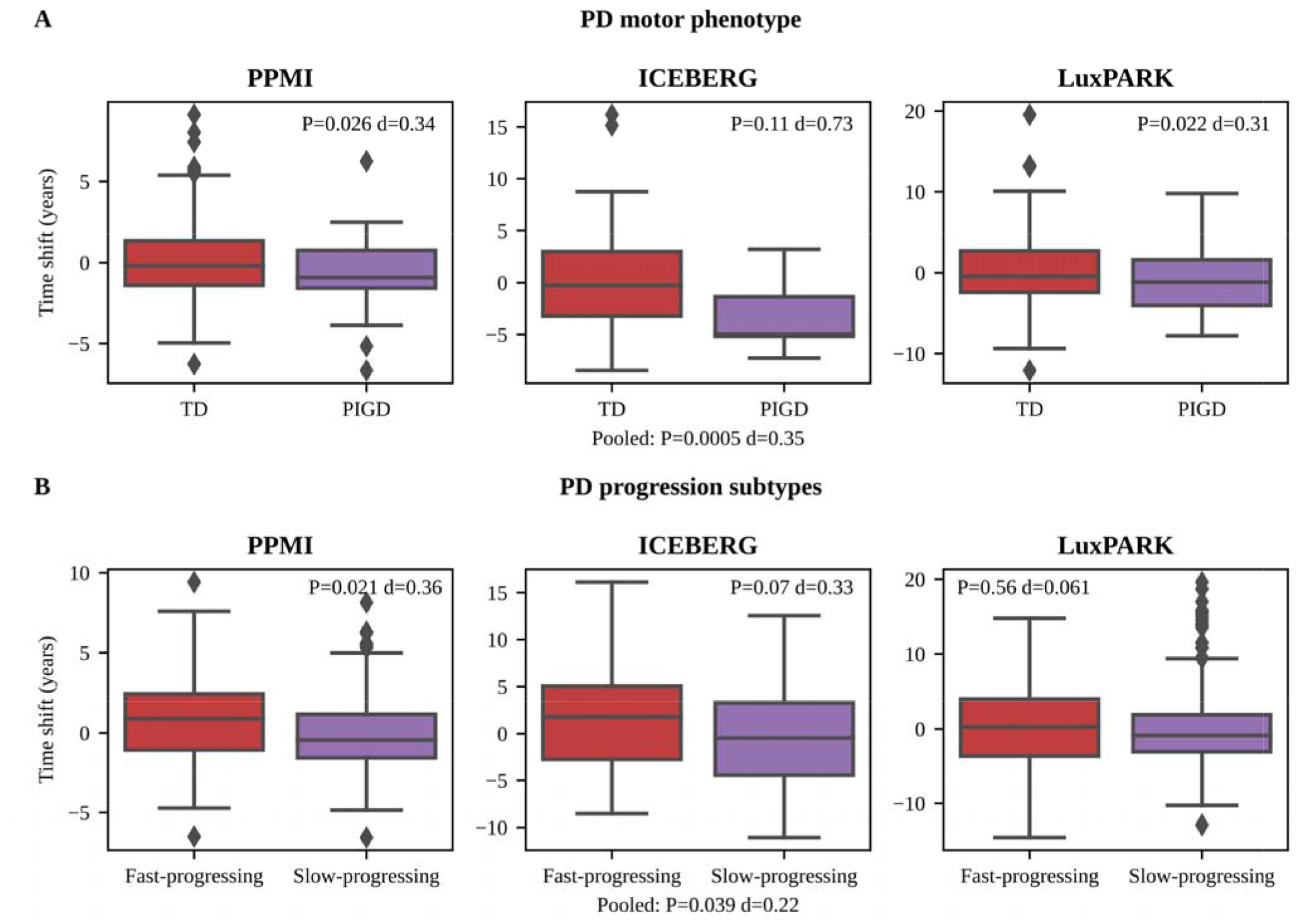
Model-derived time shift differences between motor phenotypes and PD progression subtypes. Positive model-derived time shifts indicate that PD was diagnosed later than the average patient in the corresponding cohort.

Previously, we demonstrated the existence of a fast-progressing and slow-progressing PD subtype which can potentially be biologically explained using the brain-first (slow-progressing) vs body-first (fast-progressing) model of alpha-synuclein spreading (21,25). Our pooled analysis indicates that the fast-progressing PD subtype is associated with a later-than-average PD diagnosis (d=0.22, P=0.039, Fig. **4**B, Table S4). Analyzing male and female PwPD separately, we observed no differences in model-derived time shifts between PD progression subtypes (Table S4).

Furthermore, we analyzed PwPD with PD diagnosis at younger and older age as two separate subgroups, split at the median age of PD diagnosis of 62.2 years. For the younger PwPD subgroup, associations of symptoms with a later-than-average diagnosis were mostly similar to the overall findings, except that axial and PIGD symptoms had no significant correlation with model-derived time shifts. Also, we observed no significant differences in model-derived time shifts between TD and PIGD subtype (Table S4). In contrast, PwPD in the older PD subgroup displayed fewer associations between later-than-average diagnosis and non-motor symptoms. Within this subgroup, only increased anxiety was associated with a later-than-average PD diagnosis, whereas pronounced cognitive impairment, motor symptoms, axial and PIGD symptoms, and overall disease severity correlated with an earlier-than-average PD diagnosis. Similar to the overall analysis, the TD subtype was associated with a later-than-average PD diagnosis in the older PwPD subgroup. No significant differences in model-derived time shifts were observed between PD progression subtypes in the age-specific analyses (Table S4).

Overall, we found that the fast-progressing PD subtype, more non-motor symptoms and fewer axial and PIGD symptoms were generally associated with a later-than-average PD diagnosis. However, among older PwPD, motor symptoms, overall disease severity, and cognitive impairment were also associated with an earlier-than-average PD diagnosis.

## Discussion

### Principal results

In this study, we employed a model-based approach to objectively estimate diagnostic delay and initial clinical manifestations of PD in the absence of directly observable clinical data. Analyzing longitudinal data from 1,124 PwPD from three cohorts, we identified consistent patterns of clinical and demographic characteristics associated with diagnostic delay. Older age at diagnosis, TD motor phenotype, several non-motor symptoms and a fast-progressing PD subtype were linked to a delayed PD diagnosis compared to the respective cohort averages. Furthermore, our findings highlight significant sex-specific and age-specific differences in clinical symptoms that influence diagnostic delays. In particular, non-motor symptoms played a more prominent role in delaying diagnosis for younger and female PwPD. Conversely, in older PwPD, a delay in diagnosis was observed when motor and cognitive symptoms were less pronounced.

### Estimation of diagnostic delay in PD

Previous studies assessed diagnostic delay as the interval between self-recognized onset of motor symptoms and the eventual clinical PD diagnosis (8–13,24,26,27). Adopting this traditional approach, our study observed median times between 11 and 13 months, aligning with previous findings (8– 12,24).

However, this traditional method of estimating diagnostic delay has significant limitations. The diagnostic pathway in PD can be divided into at least two distinct periods: (I) the time to symptom recognition by PwPD or relatives, and (II) the formal process of diagnosing PD (27). Previous research indicates that the formal process of diagnosing PD typically spans only a few months (8,9). This suggests that the primary contributor to diagnostic delay is the time taken for symptom recognition, a factor not captured by the traditional approach. Moreover, the traditional approach introduces a second important bias: assessing PD baseline symptoms leads to the situation that PwPD with longer diagnostic delay are likely to present a more advanced disease. This can erroneously suggest a reverse correlation between higher symptom severity and higher diagnostic delay. Lastly, structured clinical data is available solely at the point of study inclusion, rather than at the initial time of PD diagnosis. This introduces a further bias, as the progression of symptoms between diagnosis and enrollment may vary between PwPD.

Our methodology overcomes these limitations by (I) objectively estimating diagnostic delay compared to the average PD time course and (II) predicting initial manifestations at a comparable time, i.e. at the time of expected PD diagnosis. Using this model-based approach allowed us to uncover several demographic and clinical associations with diagnostic delay, which were not observable using the traditional approach.

### Characteristics leading to a delayed diagnosis

We observed highly significant and interpretable correlations of demographics and clinical symptoms with our model-derived time shifts as estimates of diagnostic delay.

Prior to our study, the relationship between age and diagnostic delay in PD was unclear (8–13). Our findings, being consistent across three cohorts, revealed a strong association of higher age with higher diagnostic delay in PD. A plausible explanation of this phenomenon could be that aged PwPD initially attribute their early symptoms to normal aging processes, leading to a delay in seeking medical consultation (26).

The impact of a positive family history of PD on diagnostic delay has also been discussed controversially (9,11,12). Two reports from Mexico suggested that a positive family history could be associated with higher diagnostic delay, possibly due to cultural factors and the absence of disease-modifying treatments deterring PwPD from seeking medical consultation (11,12). In contrast, our data collected from many different countries did not find any association between family history and diagnostic delay, thereby indicating potential regional variations in this aspect.

Furthermore, our study aligns with the majority of existing literature in finding no impact of predominant PD side or sex on diagnostic delay (8,9,11,12).

A few publications studied the impact of clinical manifestations on diagnostic delay in PD, focusing mostly on severity of motor symptoms (8–10), motor phenotype (8–12) and overall number of non-motor symptoms (6,9–11).

A previous study associated higher UPDRS III scores with increased diagnostic delay (10). We argue that this correlation may represent a reverse causation, where higher diagnostic delay resulted in more advanced PD stages at the time of diagnosis, subsequently leading to more severe motor symptoms. Contrary to this study, we have demonstrated here that more severe motor symptoms are associated with more timely diagnoses in elderly PwPD, which matches intuition.

While recent publications associated the TD motor phenotype with earlier-than-average PD diagnosis and axial symptoms with greater diagnostic delays, thereby supporting the notion of tremor as a pathognomonic and diagnosis-leading symptom for PD (9,10), our analysis presents a different perspective. We found no correlation between tremor severity and diagnostic delay. Instead, PIGD symptoms were associated with earlier-than-average diagnosis. In line with these observations, PwPD presenting with TD motor phenotype experienced higher diagnostic delays compared to those with PIGD motor phenotype. We hypothesize that the impact of PIGD symptoms on daily living, particularly their potential to cause falls, may prompt more urgent medical consultations, thereby reducing the diagnostic delay in PwPD with axial symptoms and PIGD motor phenotype. In addition, it is possible to speculate that tremor was perceived by patients as a normal age-related symptom.

While existing research focuses on the aggregated impact of non-motor symptoms on delaying PD diagnosis (6,9,10), our study adopts a more granular and systematic approach, uncovering that specifically fatigue, anxiety, depression, autonomic symptoms, pain and sleep disorders are associated with a delayed diagnosis. Correlations between non-motor symptom and diagnostic delay were even more pronounced in female and younger PwPD. This observation might be reflective of the comparatively lower incidence of PD in these demographic groups. Consequently, when non-motor symptoms predominate, patients and physicians may initially consider other conditions more likely in these populations, thereby prolonging the PD diagnostic process.

We extended the scope of this analysis beyond isolated clinical symptoms by exploring the relationship between diagnostic delay and fast vs. slow PD progression subtypes identified in previous research (21). Notably, the fast-progressing PD subtype, which could be explained biologically by the body-first concept, exhibited longer diagnostic delays. This correlation can be attributed to the initial clinical characteristics of the fast-progressing PD subtype exhibiting more non-motor (cognitive, neuropsychiatric, and autonomic) symptoms (21), which may lead to delayed PD diagnosis, as discussed in the previous paragraph.

### Limitations and future directions

By applying our LTJMM model for time shift estimations and linear mixed-effect models for prediction of initial manifestations, we assume an approximately linear progression in outcomes, without significant early-stage floor effects. However, this simplification may not fully acknowledge disease complexity (28). While incorporating exponential or sigmoid models could potentially offer a more accurate representation for these outcomes, these approaches would necessitate a more complex model, requiring increased number of patient visits for precise parameter estimations. Despite this simplification, LTJMM has been successfully applied in modeling disease progression in PD (21) and other neurodegenerative disorders (17).

While our model proficiently estimates diagnostic delay in individuals with PD relative to an average PD disease trajectory, it cannot provide specific predictions regarding the potential of earlier PD diagnosis under ideal conditions for individual PwPD. Furthermore, the individual reasons for diagnostic delays, whether due to delayed self-recognition of symptoms or a prolonged diagnostic process, remain unknown.

Based on the finding that an initial presentation with predominant non-motor symptoms delays diagnosis, it is tempting to speculate at which timepoint these patients fulfilled current diagnostic criteria for PD (29). Indeed, neurodegeneration and also non-motor symptoms often precede PD motor symptoms by several years (5,6,30). Our findings are therefore consistent with concepts that consider a diagnosis of PD that is based on biological parameters rather than classical motor symptoms (31,32). Such redefinition will become increasingly important as new disease-modifying treatments become available, aiming to stop neurodegeneration at its earliest stages. Emerging biomarkers like alpha-synuclein aggregation assays or new digital biomarkers obtained from wearables or digital assessments may also help to facilitate an earlier and more accurate PD diagnosis (33–35).

## Conclusion

Physicians and the public need to be aware that PD may manifest primarily through non-motor symptoms, especially in young and female patients. In the future, a new biological definition of PD together with emerging biomarkers may facilitate an earlier diagnosis of PD and align better with the diverse clinical manifestations of the disease described here.

## Data Availability

Relevant source code will be published at https://github.com/SCAI-BIO/PD-diagnostic-delay upon acceptance of the paper. As this study is a retrospective analysis, availability of the clinical data depends on the individual study groups (PPMI: www.ppmi-info.org, ICEBERG, LuxPARK).

