## Supplemental file for "Predictive Modeling to Uncover Parkinson’s Disease Characteristics That Delay Diagnosis"

### Clinical cohorts

#### PPMI

We analyzed 409 people with Parkinson's Disease (PwPD) from the publicly available Parkinson’s Progression Markers Initiative (PPMI, NCT04477785) with clinical visits between 2011 and 2020. All PwPD had a clinical diagnosis of Parkinson's Disease (PD) and a pathological dopamine transporter SPECT (DaTSCAN). We restricted our analysis to untreated de-novo PwPD. Therefore, we included only PwPD with clinical diagnosis not more than two years before baseline visit, Hoehn & Yahr stage 0-2 and no dopaminergic treatment at baseline visit. We further restricted our analysis to PwPD with age > 30 years and at least one additional visit as we require longitudinal information. Informed consent to data collection and sharing was obtained from all PwPD by PPMI. Ethical guidelines on human data collection were adhered to. The PPMI project was approved by the Institutional Review Board or Independent Ethics Committee of all participating sites in Europe, including Attikon University Hospital (Greece), Hospital Clinic de Barcelona and Hospital Universitario Donostia (Spain), Innsbruck University (Austria), Paracelsus-Elena-Klinic Kassel/University of Marburg (Germany), Imperial College London (UK), Pitié-Salpêtrière Hospital (France), University of Salerno (Italy), and in the USA, including Emory University, Johns Hopkins University, University of Alabama at Birmingham, PD and Movement Disorders Center of Boca Raton, Boston University, Northwestern University, University of Cincinnati, Cleveland Clinic Foundation, Baylor College of Medicine, Institute for Neurodegenerative Disorders, Columbia University Medical Center, Beth Israel Medical Center, University of Pennsylvania, Oregon Health and Science University, University of Rochester, University of California at San Diego, and University of California, San Francisco.

#### ICEBERG

We analyzed 154 PwPD from the ICEBERG cohort study (NCT02305147), an ongoing four-year observational study of PwPD with recent onset of PD conducted at the Paris Brain Institute (Institut du Cerveau-ICM, Pitié-Salpêtrière Hospital, Paris, France). Visits were performed between 2014 and 2022. PD was diagnosed according to UK Parkinson's Disease Society Brain Bank criteria and PwPD with DaTSCANs showing no dopaminergic deficit were excluded. Inclusion was restricted to disease onset not more than three years before baseline visit. We further restricted our analysis to PwPD with at least two visits as we require longitudinal information. Informed consent was obtained and ethical guidelines were adhered to. ICEBERG received approval from the local ethical committee (IRBParis VI, RCB: 2014-A00725-42).

### LuxPARK

We analyzed 561 PwPD from the Luxembourg Parkinson’s Study (LuxPARK, NCT05266872), an ongoing observational study of all disease stages PwPD from Luxembourg and the Greater Region with up to four years follow up. Visits were performed between 2015 and 2022. PD was diagnosed according to UK Parkinson's Disease Society Brain Bank criteria. We restricted our analysis to PwPD with at least two visits as we require longitudinal information. Informed consent was obtained and ethical guidelines were adhered to. LuxPARK was approved by the National Ethics Board in Luxembourg (CNER Ref: 201407/13).

### Latent time joint mixed-effects model

We modeled disease progression in each cohort as a linear process using a latent time joint mixed-effects model (LTJMM) as proposed from Li et. al.[1]

$$y_{ijk} = x_i \beta_k + \gamma_k (t_{ijk} + \delta_i) + \alpha_{0ik} + \alpha_{1ik} t_{ijk} + \epsilon_{ijk}$$

Thereby, we denote $y_{ijk}$ as outcome k observed at measurement j for an individual i. We account for age and sex differences by including age at diagnosis and sex as covariates $x_i$ into the model with $\beta_k$ as corresponding coefficient shared across all individuals. The coefficient $\gamma_k$ represents the mean slope of the cohort for each outcome k and is thereby shared across all individuals. We use the time since diagnosis as $t_{ijk}$ and shift all measurements of an individual by a PwPD specific time shift $\delta_i$ shared across all outcomes. Additionally, we include random intercepts $\alpha_{0ik}$ and random slopes $\alpha_{1ik}$ for each individual and outcome. As usual, measurement errors $\varepsilon_{ijk}$ and time shifts $\delta_i$ are both assumed to be drawn from normal distributions with a mean of zero. Random intercepts and slopes follow a multivariate normal distribution with mean of zero. Fitting was performed using a Markov chain Monte Carlo (MCMC) algorithm with 4 chains, 25000 iterations and 12500 warm up steps. Analyses were performed using the R packages ltjmm[2] and rstan.[3]

Unified Parkinson's Disease Rating Scale (UPDRS) I-IV, Postural Instability and Gait Dysfunction score (PIGD), Montreal Cognitive Assessment (MoCA) and Scales for Outcomes in Parkinson's Disease-Autonomic Dysfunction (SCOPA) were used as outcomes and min-max-normalized on the theoretical range of the scores. MoCA scores were inverted to ensure positive slopes for all outcomes.

Convergence of MCMCs and normal distribution of parameter estimates were inspected manually. In addition, $\hat{R}$ statistics were calculated and ensured to be below 1.05.

To visualize and validate the effect of aligning PwPD on a common timescale, we inspected the distributions of Hoehn & Yahr (H&Y) stages which were not used for fitting the LTJMM model. Thereby, we observed a clearer separation of H&Y stages after applying LTJMM to the data and a stronger correlation between H&Y stages and the timescale (Fig. S3).

Further, we inspected the accuracy of our LTJMM approach in predicting outcomes at the next visit. Therefore, we re-trained LTJMM, but excluded the last measurement of all outcomes. Using this LTJMM model, we predicted these last measurements of all outcomes and calculated the coefficient of determination ($R^2$) for these predictions. Thereby, we obtained reasonable $R^2$ values: 55% (PPMI), 53% (ICEBERG), 61% (LuxPARK).

To analyze (I) the correlation of time shifts with age at diagnosis and (II) the time shift differences between female and male PwPD, we used a simplified LTJMM model without sex and age at diagnosis as covariates.

#### Estimating clinical manifestation at the time when PD is diagnosed typically

Clinical symptoms are only reported at the time of study visits and not available at the time of PD diagnosis. Therefore, we estimated the initial clinical manifestations at the time of PD diagnosis on the common disease timescale, i.e. at the time where $t_i + \delta_i = 0$. This was achieved by applying the following steps to every outcome listed in Table S1: (I) aligning all measurements on the common disease timescale, (II) fitting linear, ordinal or binary mixed effect models on the measurement time series based on the scale of the outcome, (III) predicting the initial manifestation, i.e. the outcome at the time where $t_i + \delta_i = 0$ on the common disease timescale. We restricted our analysis to outcomes with at least 2 measurements per PwPD as we require longitudinal data. Furthermore, we restricted our analysis to outcomes with data available for at least 30 PwPD.

To validate our model predictions, we compared the coefficient of determination with a null model for all three cohorts. More precisely, linear, ordinal and binary mixed effect models were fitted by leaving out the value of the first visit. $R^2$ values of the first-visit predictions were calculated and compared with a null-model using the value of the second visit. For the null model, only visits being at least one year apart from the first visit were taken into account. A Wilcoxon signed rank-test was used to compare $R^2$ values of the null model with the statistical models and indicated that the models outperformed the null model in all cohorts ($P<0.0001$, Fig. S4).

The statistical models were fitted using the R packages lme4[4] and ordinal.[5]

### Symptom domain comparisons

To assess the influence of clinical manifestations on the time of PD diagnosis, we analyzed the correlation of (I) patient-reported time to diagnosis with baseline clinical manifestations and (II) model-derived time shifts estimated by the LTJMM and initial clinical manifestations estimated by the statistical models.

Correlations between outcomes and the time variable (i.e., time to diagnosis or model-derived time shifts) were calculated as Pearson, Kendall Tau-b and point-biserial correlation, depending on the scale of the outcome.

To allow a more comprehensive analysis and validation of the variety of outcomes captured across the three cohorts, we grouped 66 outcomes (including single questions, scores and sub-scores from questionnaires and clinical assessments) into 17 symptom domains (Table S1). The choice of the 17 symptom domains represents a trade-off between capturing most clinically relevant motor and non-motor symptoms and which outcomes had been assessed in the three cohorts.

To assess the correlation of clinical outcomes with the time variable in the defined symptom domains, we conducted a three-level meta-analysis with random effects for each symptom domain. Therefore, we first calculated an overall regression coefficient across all outcomes of a symptom domain per cohort. Subsequently, we calculated an overall regression coefficient estimate of the symptom domain across the three cohorts (see forest plots at end of the supplement). P-values and 95% confidence intervals (CI) were corrected for multiple testing across the 17 symptom domains using the Benjamini-Hochberg procedure.[6]

Meta analyses were performed using the R-package meta.[7]

### Supplementary figures and tables

| Symptom domain | Outcome | Definition/calculation of the outcome |
|---|---|---|
| Anxiety | NMSQ Anxiety | NMSQ item 17 |
| | HADS anxiety | HADS anxiety sub-score |
| | STA | STA sum score |
| | PDQ39 Anxiety | PDQ39 item 21 |
| | UPDRS I Anxiety | UPDRS I item 4 |
| Apathy | DAS | DAS sum score |
| | SAS | SAS sum score |
| | UPDRS I Apathy | UPDRS I item 5 |
| Autonomic symptoms | NMSQ Autonomic | NMSQ sum of items 4, 5, 6, 7, 8, 9, 19, 20, 28 |
| | SCOPA-AUT | SCOPA sum score |
| | UPDRS I Autonomic | UPDRS I sum of items 10, 11, 12 |
| Overall Cognition | MATTIS | MATTIS sum score |
| | MMSE | MMSE sum score |
| | MoCA | MoCA sum score |
| | SIQCDE | Short IQCODE score sum score |
| | FAB | FAB sum score |
| | PDQ39 Cognition | PDQ39 sum of items 31, 32 |
| | UPDRS I Cognition | UPDRS I item 1 |
| | NMSQ Cognition | NMSQ sum of items 12, 15 |
| Depression | BDI | BDI sum score |
| | GDS | GDS sum score |
| | HADS depression | HADS depression sub-score |
| | PDQ39 Depression | PDQ39 sum of items 17, 18, 19, 20, 22 |
| | NMSQ Depression | NMSQ sum of items 13, 16 |
| | UPDRS I Depression | UPDRS 1 item 3 |
| Fatigue | UPDRS I Fatigue | UPDRS 1 item 13 |
| Hallucinations | NMSQ Hallucination | NMSQ sum of items 14, 30 |
| | UPDRS I Hallucinations | UPDRS 1 item 2 |
| Impulsivity | QUIP | QUIP sum score |
| | QUIP-RS | QUIPRS sum score |
| Motor symptoms (overall) | PDQ39 ADL | PDQ39 ADL sub-score |
| | Pegboard | PEGBoard sum of: average of left hand, right hand and both hands |
| | UPDRS II | UPDRS II sum score |
| | UPDRS III off | UPDRS III sum score (OFF only) |
| | UPDRS IV | UPDRS IV sum score |
| Non motor symptoms (overall) | NMSQ | NMSQ sum score |
| | UPDRS I | UPDRS I sum score |
| Overall disease severity | UPDRS I-III off | UPDRS I, II, III sum (OFF only) |
| | FAQ | FAQ sum score |

| Symptom domain | Outcome | Definition/calculation of the outcome |
|---|---|---|
| | PDQ39 | PDQ39 sum score |
| | SEADL | SEADL score |
| | H&Y | Hoehn & Yahr |
| | CGIS | CGI-S score |
| Pain | NMSQ Pain | NMSQ item 10 |
| | PDQ39 Pain | PDQ39 sum of items 37, 38 |
| | UPDRS I Pain | UPDRS 1 item 9 |
| Axial & PIGD symptoms | UPDRS III axial off | UPDRS III axial score (OFF only) |
| | FOGAC | FOGAC sum score |
| | FOGQ | FOGQ sum score |
| | GABS Examination | GABS sum of items 8-24 |
| | GABS Questionnaire | GABS sum of items 1-7 |
| | NFOGQ | NFOGQ sum score |
| | PDQ39 Mobility | PDQ39 mobility sub-score |
| | PIGD off | PIGD score (OFF only) |
| | TUG | Timed Up and Go time |
| Sleep (general) | ESS | ESS sum score |
| | PDSS | PDSS sum score |
| | UPDRS I Sleep | UPDRS I sum of items 7, 8 |
| | NMSQ Sleep | NMSQ sub of items 22, 23 |
| RBD Sleep | RBD-HK | RBD-HK sum score |
| | RBD-SQ | RBD-SQ sum score |
| | NMSQ RBD | NMSQ sum of items 24, 25 |
| Smell | NMSQ Smell | NMSQ item 2 |
| | Sniffin Test | Sniffin Test score |
| | UPSIT | UPSIT sum score |
| Tremor | TD off | TD score (OFF only) |

***Table S1: Construction of symptom domains***

*Abbreviations: BDI: Beck Depression Inventory, CGIS: Clinical Global Impression-Severity, DAS: Dimensional Apathy Scale, ESS: Epworth Sleepiness Scale, FAB: Frontal Assessment Battery, FAQ: Functional Activities Questionnaire, FOGAC: Freezing of Gait AC, FOGQ: Freezing of Gait Questionnaire, GABS: Clinical Gait and Balance Scale, GDS: Geriatric Depression Scale, H&Y: Hoehn & Yahr scale, HADS: Hospital Anxiety and Depression Scale, MATTIS: Mattis Dementia Rating Scale, MMSE: Mini Mental Status Examination, MOCA: Montreal Cognitive Assessment, NFOGQ: New Freezing of Gait Questionnaire, NMSQ: Non-Motor Symptoms Questionnaire, PDQ39: Parkinson's Disease Questionnaire-39, PDSS: Parkinson's Disease Sleep Scale, PIGD: Postural Instability and Gait Disorder score, QUIP: Questionnaire for Impulsive-Compulsive Disorders, QUIP-RS: QUIP-Rating Scale, RBD-HK: REM Sleep Behavior Disorder Questionnaire-Hong Kong, RBD-SQ: REM Sleep Behavior Disorder Screening Questionnaire, SAS: Starkstein Apathy Scale, SCOPA-AUT: Scales for Outcomes in Parkinson's Disease-Autonomic Dysfunction, SEADL: Schwab and England Activities of Daily Living Scale, SIQCDE: Short Informant Questionnaire on Cognitive Decline in the Elderly, STA: State-Trait Anxiety Inventory, TD: Tremor Dominance Score, TUG: Timed Up and Go, UPDRS: MDS-Unified Parkinson's Disease Rating Scale, UPSIT: University of Pennsylvania Smell Identification Test*

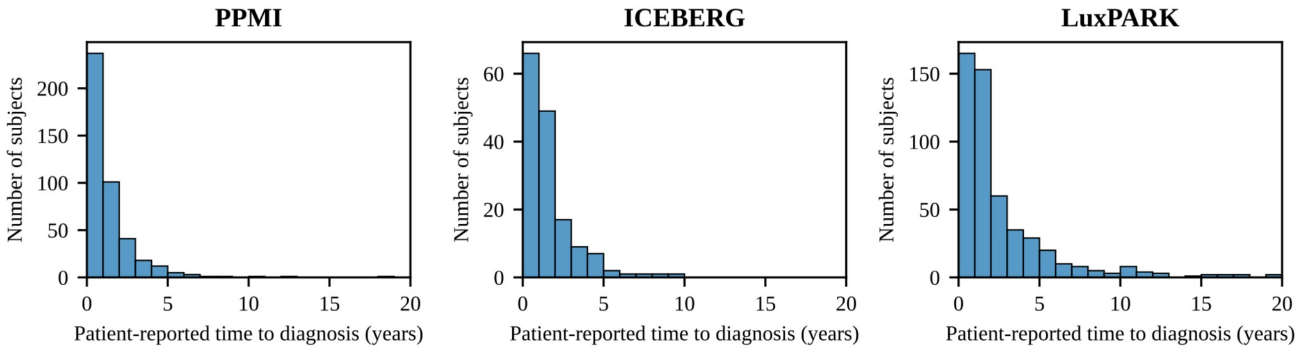

***Figure S1: Distribution of patient-reported time to diagnosis across cohorts***
*The histograms depict the distribution of patient-reported time to diagnosis for PPMI, ICEBERG and LuxPARK cohorts. Patient-reported time to diagnosis was defined as the time span between occurrence of first motor symptom and PD diagnosis.*

| | PPMI | ICEBERG | LuxPARK |
|---|---|---|---|
| **Age at diagnosis** | ρ=-0.076 (P=0.12) | ρ=0.0017 (P=0.98) | ρ=0.035 (P=0.42) |
| **Sex** (female/male) | d=0.0053 (P=0.37) | d=0.20 (P=0.19) | d=0.0081 (P=0.90) |
| **Family history of PD** (no/yes) | d=-0.11 (P=0.31) | d=0.16 (P=0.93) | d=0.045 (P=0.87) |
| **Motor phenotype** (TD/PIGD) | d=0.24 (P=0.20) | d=0.15 (P=0.83) | d=0.21 (P=0.20) |
| **Predominant side** (left/right) | d=0.01 (P=0.46) | d=0.16 (P=0.13) | d=0.084 (P=0.91) |

***Table S2: Associations of demographic and clinical characteristics with patient-reported time to diagnosis***

*The relationships between age at PD diagnosis and patient-reported time to diagnosis were assessed using Pearson correlation with corresponding correlation coefficients and p-values being reported. Regarding sex, family history of PD, motor phenotype and predominant side, patient-reported times to diagnosis were compared between subgroups using Mann-Whitney U tests. Corresponding p-values and Cohen’s D are reported. Positive Cohen’s D values indicate a higher patient-reported time to diagnosis for female PwPD, negative family history, PwPD with TD phenotype and PwPD with left predominant side. Pooled estimates were calculated from meta-analyses across the three cohorts.*

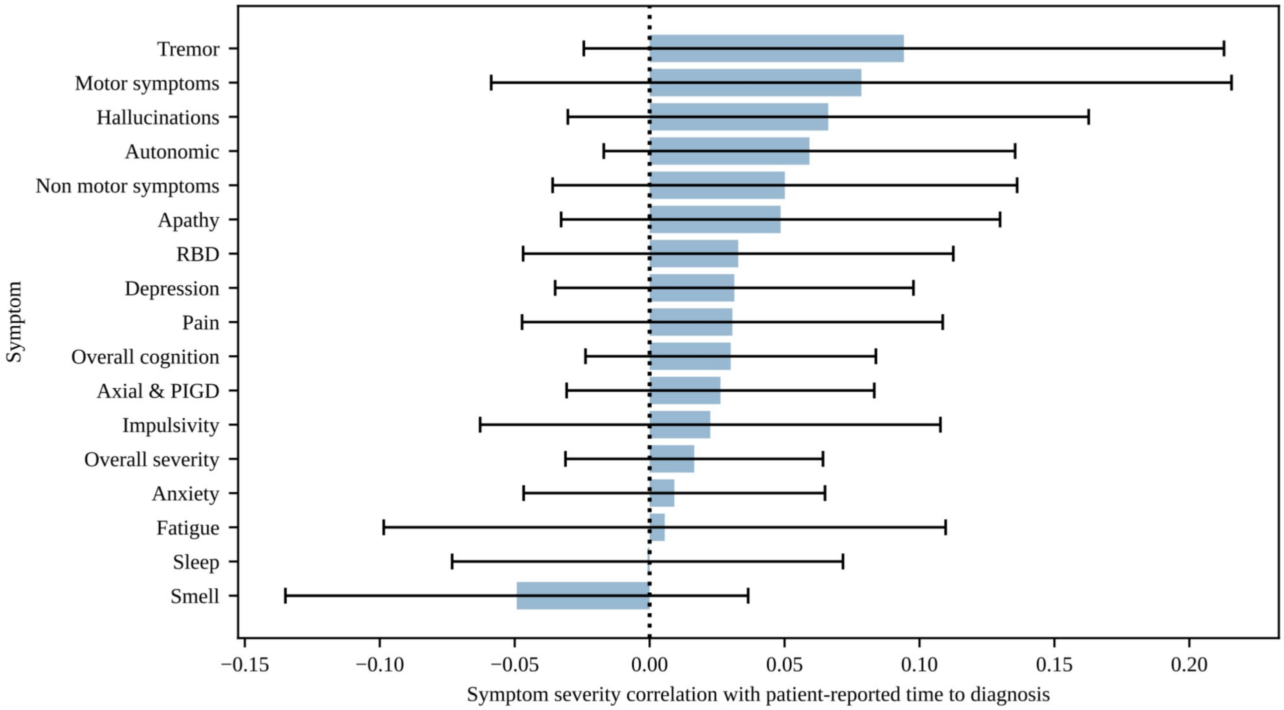

***Figure S2 Correlations of baseline clinical characteristics with patient-reported time to diagnosis***
*The figure depicts the correlations between the severity of different symptom domains at baseline visit and the patient-reported time to diagnosis. Positive correlation coefficients mean that increased symptom severity is associated with a longer patient-reported time to diagnosis. The presented correlation coefficients are pooled estimates derived from several clinical scores from the PPMI and ICEBERG cohort. The LuxPARK cohort was not included into this analysis as it included also advanced disease stage PwPD at baseline visit. Confidence intervals were corrected for multiple testing. P-values and correlation coefficients are also reported in Table S3.*
*Abbreviations: RBD: REM behavior sleep disorder, PIGD: postural instability and gait disturbance*

| Symptom | P value | Correlation coefficient |
|---|---|---|
| **Anxiety** | 0.84 | 0.01 |
| **Apathy** | 0.44 | 0.05 |
| **Autonomic** | 0.27 | 0.06 |
| **Axial & PIGD** | 0.54 | 0.03 |
| **Depression** | 0.54 | 0.03 |
| **Fatigue** | 0.97 | 0.01 |
| **Hallucinations** | 0.41 | 0.07 |
| **Impulsivity** | 0.72 | 0.02 |
| **Motor symptoms** | 0.44 | 0.08 |
| **Non motor symptoms** | 0.44 | 0.05 |
| **Overall cognition** | 0.44 | 0.03 |
| **Overall severity** | 0.62 | 0.02 |
| **Pain** | 0.58 | 0.03 |
| **RBD** | 0.58 | 0.03 |
| **Sleep** | 0.98 | 0.00 |
| **Smell** | 0.44 | -0.05 |
| **Tremor** | 0.27 | 0.09 |

***Table S3: Correlations of baseline clinical characteristics with patient-reported time to diagnosis***

*The table reports the correlations of baseline clinical characteristics with patient-reported time to diagnosis. Positive correlation coefficients mean that increased symptom severity is associated with a longer patient-reported time to diagnosis. The presented correlation coefficients are pooled estimates derived from several clinical scores from the PPMI and ICEBERG cohort. The LuxPARK cohort was not included into this analysis as it included also advanced disease stage PwPD at baseline visit. Corresponding p-values were corrected for multiple testing.*

*Abbreviations: RBD: REM behavior sleep disorder, PIGD: postural instability and gait disturbance*

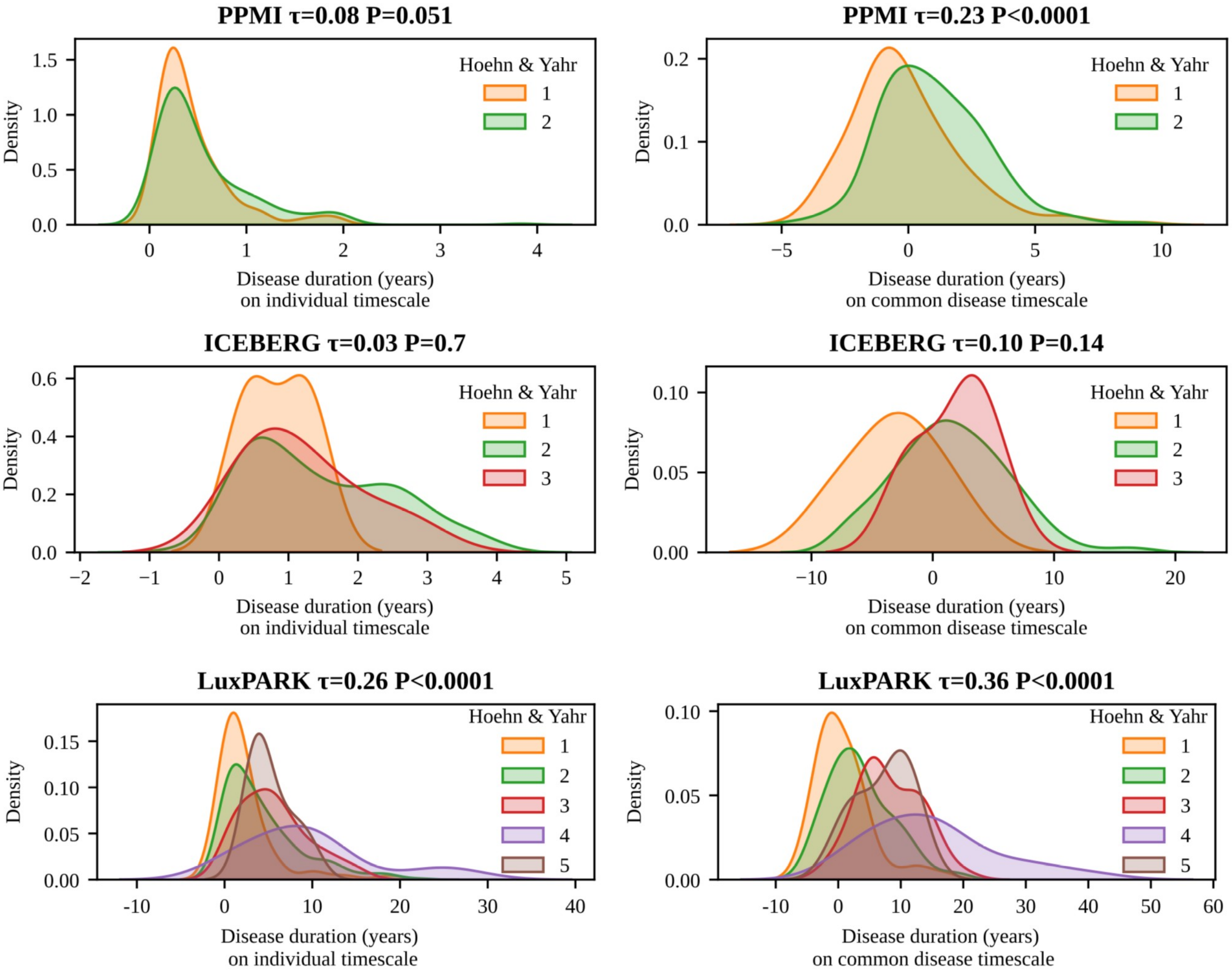

***Figure S3: Effect of time-aligning PwPD on distributions of H&Y stages***

*H&Y baseline distributions from PPMI, ICEBERG and LuxPARK are depicted as kernel density estimation plots. On the left side, H&Y stages are plotted against the original timescale. On the right side, H&Y stages are plotted against the common disease timescale calculated from the LTJMM. Correlation of H&Y stages with the timescale are reported by Kendall tau-b correlation coefficients and corresponding p-values. Thereby, stronger correlations are observed after aligning PwPD on the common disease timescale.*

*Abbreviations: H&Y: Hoehn&Yahr, LTJMM: latent time joint mixed-effects model*

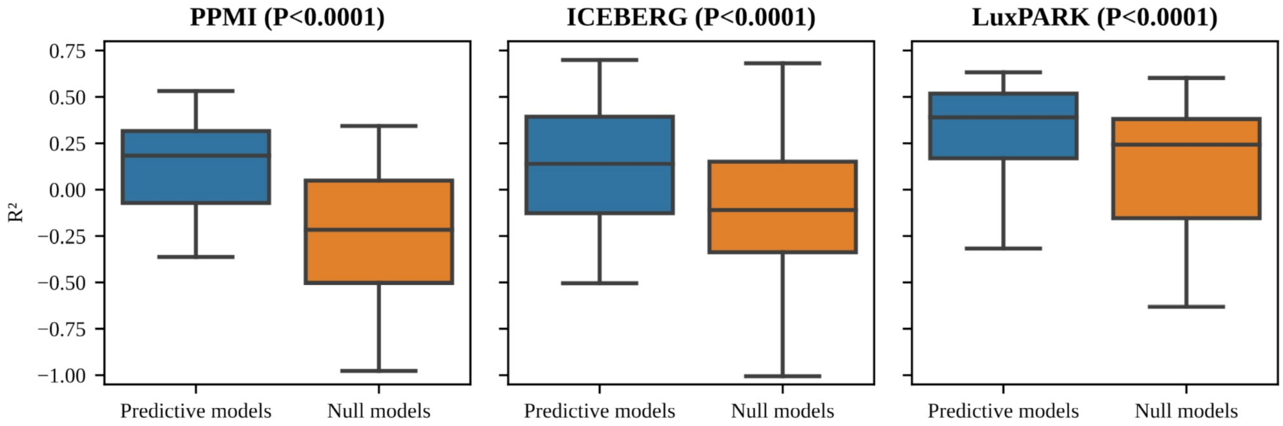

***Figure S4: Predictive performance of statistical models***

*Predictive performance of statistical models compared to a null model for the three cohorts. Linear, ordinal and binary mixed effect models were fitted to the longitudinal data of the outcomes listed in Table S1, thereby leaving out the value of the first visit.* $R^2$ *values of the first-visit predictions were calculated and compared with a null-model using the value of the second visit. For the null model, only visits being at least one year apart from the first visit were taken into account. Corresponding p-values were calculated using a Wilcoxon signed-rank test.*

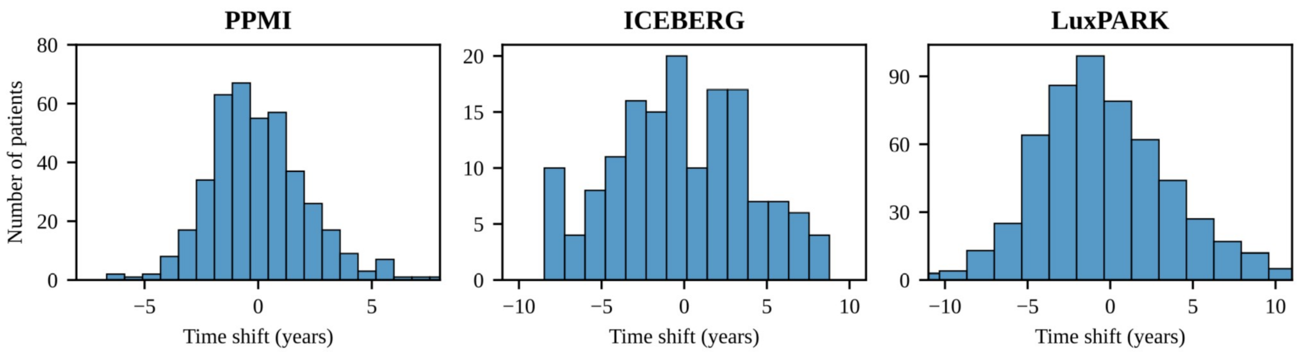

***Figure S5 Distribution of model-derived time shifts across cohorts***
*The histograms depict the variation in model-derived time shifts for PPMI, ICEBERG and LuxPARK cohorts in relation to an average PwPD. Positive model-derived time shifts indicate that PD was diagnosed later than average.*
*Abbreviations: LTJMM: latent time joint mixed-effects model.*

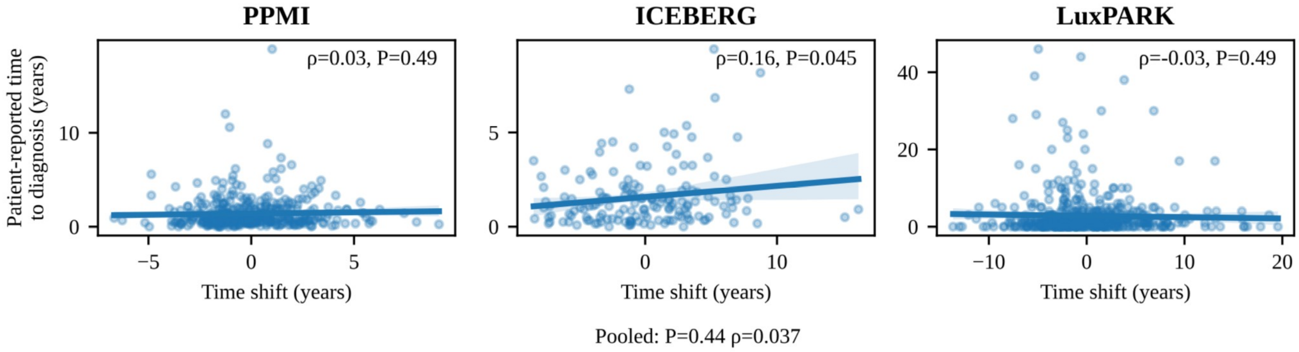

***Figure S6: Correlation of patient-reported time to diagnosis with model-derived time shifts***

*Positive model-derived time shifts indicate that PD was diagnosed later than for an average PwPD in the corresponding cohort. Pearson correlation coefficients and corresponding p-values are shown for the correlations between patient-reported time to diagnosis and model-derived time shifts.*

| | | PPMI | ICEBERG | LuxPARK | Pooled estimate |
|---|---|---|---|---|---|
| **Predicted age at diagnosis** | | **ρ=0.43<br>(P<0.0001)** | **ρ=0.74<br>(P<0.0001)** | **ρ=0.47<br>(P<0.0001)** | **ρ=0.56<br>(P<0.0001)** |
| | **female** | **ρ=0.25<br>(P=0.0037)** | **ρ=0.68<br>(P<0.0001)** | **ρ=0.46<br>(P<0.0001)** | **ρ=0.48<br>(P=0.0015)** |
| | **male** | **ρ=0.51<br>(P<0.0001)** | **ρ=0.77<br>(P<0.0001)** | **ρ=0.48<br>(P<0.0001)** | **ρ=0.60<br>(P<0.0001)** |
| **Sex**<br>(female/male) | | d=0.099<br>(P=0.34) | d=0.13<br>(P=0.45) | d=-0.079<br>(P=0.38) | d=0.022<br>(P=0.76) |
| **Family history of PD**<br>(no/yes) | | d=-0.079<br>(P=0.49) | d=0.12<br>(P=0.52) | d=0.041<br>(P=0.67) | d=-0.0084<br>(P=0.90) |
| **Predicted motor phenotype**<br>(TD/PIGD) | | **d=0.34<br>(P=0.026)** | d=0.73<br>(P=0.11) | **d=0.31<br>(P=0.022)** | **d=0.35<br>(P=0.0005)** |
| | **young (<62.2 years)** | d=0.13<br>(P=0.58) | d=0.86<br>(P=0.15) | .. | d=0.29<br>(P=0.34) |
| | **old (>= 62.2 years)** | **d=0.53<br>(P=0.013)** | .. | **d=0.31<br>(P=0.022)** | **d=0.38<br>(P=0.0010)** |
| | **female** | d=0.29<br>(P=0.24) | d=0.88<br>(P=0.15) | d=0.38<br>(P=0.11) | **d=0.38<br>(P=0.02)** |
| | **male** | d=0.38<br>(P=0.059) | .. | d=0.28<br>(P=0.094) | **d=0.32<br>(P=0.012)** |
| **Predominant side**<br>(left/right) | | d=-0.099<br>(P=0.32) | d=-0.015<br>(P=0.93) | d=0.18<br>(P=0.19) | d=0.0069<br>(P=0.94) |
| **Progression subtype**<br>(fast-progressing/slow-progressing) | | **d=0.36<br>(P=0.021)** | d=0.33<br>(P=0.07) | d=0.061<br>(P=0.56) | **d=0.22<br>(P=0.039)** |
| | **young (<62.2 years)** | d=0.23<br>(P=0.32) | **d=0.86<br>(P=0.0015)** | d=0.052<br>(P=0.78) | d=0.34<br>(P=0.14) |
| | **old (>= 62.2 years)** | d=0.3<br>(P=0.12) | d=-0.26<br>(P=0.33) | d=0.019<br>(P=0.89) | d=0.066<br>(P=0.62) |
| | **female** | d=-0.061<br>(P=0.85) | d=0.20<br>(P=0.54) | d=0.28<br>(P=0.11) | d=0.17<br>(P=0.20) |
| | **male** | **d=0.54<br>(P=0.00043)** | d=0.37<br>(P=0.096) | d=-0.029<br>(P=0.82) | d=0.28<br>(P=0.14) |

***Table S4: Associations of demographic and clinical characteristics with model-derived time shifts***
*The relationships between predicted age at PD diagnosis and model-derived time shifts were assessed using Pearson correlation with corresponding p-values being reported. Regarding sex, family history of PD, predicted motor phenotype, predominant side and progression subtype, model-derived time shifts were compared between subgroups using t-tests or Welch's test depending on the distribution of the data. Corresponding p-values and Cohen's D are shown. Thereby, positive Cohen's D values indicate a higher model-derived time shift for female PwPD, negative family history, PwPD with predicted TD phenotype, PwPD with left predominant side and the fast-progressing PD subtype. Pooled estimates were calculated from meta-analyses across the three cohorts. Analyses of sex and predicted age at diagnosis were carried out using the LTJMM model without age at diagnosis and sex as covariates. Analyses of family history, motor phenotype, progression subtype and predominant side associations were carried out using the LTJMM model including age at diagnosis and sex as covariates. Significant results are indicated in bold. Subgroup analyses were carried out for characteristics with a significant pooled effect. Two dots (..) indicate that not enough PwPD were available for at least one motor phenotype in the corresponding cohort subgroup.*

| Symptom domain | All PwPD | Male PwPD | Female PwPD | Early onset PwPD (Age at diagnosis <= 62.2 years) | Late onset PwPD (Age at diagnosis > 62.2 years) |
|---|---|---|---|---|---|
| **Anxiety** | **0.13 (P=0.0043)** | **0.12 (P=0.005)** | **0.14 (P=0.019)** | **0.17 (P=0.00036)** | **0.11 (P=0.025)** |
| **Apathy** | 0.02 (P=0.32) | 0.03 (P=0.41) | 0.0 (P=0.99) | 0.05 (P=0.23) | -0.04 (P=0.36) |
| **Autonomic** | **0.1 (P=0.0019)** | **0.1 (P=0.014)** | **0.11 (P=0.0051)** | **0.14 (P=0.015)** | 0.05 (P=0.098) |
| **Overall cognition** | 0.0 (P=0.88) | 0.0 (P=0.95) | 0.01 (P=0.92) | 0.03 (P=0.42) | **-0.07 (P=0.022)** |
| **Depression** | **0.1 (P=0.0004)** | **0.11 (P=0.0005)** | **0.09 (P=0.011)** | **0.1 (P=0.025)** | 0.05 (P=0.23) |
| **Fatigue** | **0.13 (P=0.012)** | 0.11 (P=0.084) | 0.21 (P=0.13) | 0.2 (P=0.088) | 0.08 (P=0.36) |
| **Hallucinations** | -0.05 (P=0.092) | -0.02 (P=0.7) | -0.07 (P=0.26) | -0.06 (P=0.088) | -0.07 (P=0.19) |
| **Impulsivity** | 0.07 (P=0.12) | 0.02 (P=0.78) | **0.19 (P=0.019)** | 0.11 (P=0.097) | 0.07 (P=0.36) |
| **Motor symptoms** | -0.05 (P=0.23) | -0.05 (P=0.41) | -0.04 (P=0.3) | 0.0 (P=0.97) | **-0.13 (P=0.00014)** |
| **Non motor symptoms** | **0.15 (P=0.0006)** | **0.14 (P=0.0082)** | **0.14 (P=0.0077)** | **0.23 (P=0.0007)** | 0.04 (P=0.36) |
| **Overall severity** | -0.04 (P=0.28) | -0.055 (P=0.3) | -0.07 (P=0.14) | 0.01 (P=0.94) | **-0.14 (P<0.0001)** |
| **Pain** | **0.08 (P=0.0085)** | 0.07 (P=0.21) | **0.11 (P=0.019)** | **0.12 (P=0.007)** | 0.08 (P=0.12) |
| **Axial & PIGD** | **-0.12 (P=0.0004)** | **-0.14 (P<0.0001)** | **-0.13 (P=0.011)** | -0.06 (P=0.069) | **-0.19 (P<0.0001)** |
| **Sleep** | **0.06 (P=0.0043)** | 0.04 (P=0.21) | **0.12 (P=0.0029)** | **0.09 (P=0.0017)** | 0.01 (P=0.77) |
| **RBD** | 0.06 (P=0.28) | 0.07 (P=0.27) | 0.05 (P=0.42) | 0.03 (P=0.82) | 0.04 (P=0.47) |
| **Smell** | -0.03 (P=0.79) | -0.05 (P=0.68) | -0.06 (P=0.57) | -0.08 (P=0.42) | -0.04 (P=0.74) |
| **Tremor** | -0.03 (P=0.75) | 0.03 (P=0.78) | -0.12 (P=0.13) | -0.05 (P=0.48) | -0.04 (P=0.53) |

***Table S5: Correlation between initial clinical characteristics and model-derived time shifts in PD for subgroups***

*The table reports the correlations of estimated initial clinical characteristics at the point of a typical PD diagnosis with model-derived time shifts. Positive correlation coefficients mean that increased symptom severity is associated with a PD diagnosis later than average. The presented correlation coefficients are pooled estimates derived from the three cohorts and several clinical scores. Corresponding p-values were corrected for multiple testing. Significant correlations are indicated bold. Results are shown for the overall PD cohorts, a sex-specific subgroup analysis and a subgroup analysis based on a median split of age at diagnosis.*

*Abbreviations: RBD: REM behavior sleep disorder, PIGD: postural instability and gait disturbance*

### Forest plots for correlation of baseline symptom domains with patient-reported time to diagnosis

### Forest plots for correlation of initial symptom domains with model-derived time shifts

### Acknowledgment

#### Parkinson’s Progression Markers Initiative

Data used in the preparation of this article were obtained from the Parkinson’s Progression Markers Initiative (PPMI) database www.ppmi-info.org/data. PPMI—a public-private partnership – is funded by the Michael J. Fox Foundation for Parkinson’s Research and funding partners. A list of names of all the PPMI funding partners can be found at www.ppmi-info.org/about-ppmi/who-we-are/study-sponsors/.

#### ICEBERG study group

**Steering committee:** Marie Vidailhet, MD, PhD, (Pitié-Salpêtrière Hospital, Paris, principal investigator of ICEBERG), Jean-Christophe Corvol, MD, PhD (Pitié-Salpêtrière Hospital, Paris, scientific lead), Isabelle Arnulf, MD, PhD (Pitié-Salpêtrière Hospital, Paris, member of the steering committee), Stéphane Lehericy, MD, PhD (Pitié-Salpêtrière Hospital, Paris, member of the steering committee);

**Clinical data:** Marie Vidailhet, MD, PhD, (Pitié-Salpêtrière Hospital, Paris, coordination), Graziella Mangone, MD, PhD (Pitié-Salpêtrière Hospital, Paris, co-coordination), Jean-Christophe Corvol, MD, PhD (Pitié-Salpêtrière Hospital, Paris), Isabelle Arnulf, MD, PhD (Pitié-Salpêtrière Hospital, Paris), Sara Sambin, MD (Pitié-Salpêtrière Hospital, Paris), Poornima Menon, MD (Pitié-Salpêtrière Hospital, Paris, Jonas Ihle, MD (Pitié-Salpêtrière Hospital, Paris), Caroline Weill, MD, (Pitié-Salpêtrière Hospital, Paris), David Grabli, MD, PhD (Pitié-Salpêtrière Hospital, Paris); Florence Cormier-Dequaire, MD (Pitié-Salpêtrière Hospital, Paris); Louise Laure Mariani, MD, PhD (Pitié-Salpêtrière Hospital, Paris), Bertrand Degos, MD, PhD (Avicenne Hospital, Bobigny);

**Neuropsychological data:** Richard Levy, MD (Pitié-Salpêtrière Hospital, Paris, coordination), Fanny Pineau, MS (Pitié-Salpêtrière Hospital, Paris, neuropsychologist), Julie Socha, MS (Pitié-Salpêtrière Hospital, Paris, neuropsychologist), Eve Benchetrit, MS (La Timone Hospital, Marseille, neuropsychologist), Virginie Czernecki, MS (Pitié-Salpêtrière Hospital, Paris, neuropsychologist), Marie-Alexandrine, MS (Pitié-Salpêtrière Hospital, Paris, neuropsychologist);

**Eye movement:** Sophie Rivaud-Pechoux, PhD (ICM, Paris, coordination); Elodie Hainque, MD, PhD (Pitié-Salpêtrière Hospital, Paris);

**Sleep assessment:** Isabelle Arnulf, MD, PhD (Pitié-Salpêtrière Hospital, Paris, coordination), Smaranda Leu Semenescu, MD (Pitié-Salpêtrière Hospital, Paris), Pauline Dodet, MD (Pitié-Salpêtrière Hospital, Paris);

**Genetic data:** Jean-Christophe Corvol, MD, PhD (Pitié-Salpêtrière Hospital, Paris, coordination), Graziella Mangone, MD, PhD (Pitié-Salpêtrière Hospital, Paris, co-coordination), Samir Bekadar, MS

(Pitié-Salpêtrière Hospital, Paris, biostatistician), Alexis Brice, MD (ICM, Pitié-Salpêtrière Hospital, Paris), Suzanne Lesage, PhD (INSERM, ICM, Paris, genetic analyses);

**Metabolomics:** Fanny Mochel, MD, PhD (Pitié-Salpêtrière Hospital, Paris, coordination), Farid Ichou, PhD (ICAN, Pitié-Salpêtrière Hospital, Paris), Vincent Perlbarg, PhD, Pierre and Marie Curie University), Benoit Colsch, PhD (CEA, Saclay), Arthur Tenenhaus, PhD (Supelec, Gif-sur-Yvette, data integration);

**Brain MRI data:** Stéphane Lehericy, MD, PhD (Pitié-Salpêtrière Hospital, Paris, coordination), Rahul Gaurav, MS, (Pitié-Salpêtrière Hospital, Paris, data analysis), Nadya Pyatigorskaya, MD, PhD, (Pitié-Salpêtrière Hospital, Paris, data analysis); Lydia Yahia-Cherif, PhD (ICM, Paris, Biostatistics), Romain Valabregue, PhD (ICM, Paris, data analysis), Cécile Galléa, PhD (ICM, Paris);

**Datscan imaging data:** Marie-Odile Habert, MCU-PH (Pitié-Salpêtrière Hospital, Paris, coordination);

**Voice recording:** Dijana Petrovska, PhD (Telecom Sud Paris, Evry, coordination), Laetitia Jeancolas, MS (Telecom Sud Paris, Evry);

**Study management:** Alizé Chalançon (Pitié-Salpêtrière Hospital, Paris, Project manager), Carole Dongmo-Kenfack (Pitié-Salpêtrière Hospital, Paris, clinical research assistant); Christelle Laganot (Pitié-Salpêtrière Hospital, Paris, clinical research assistant), Valentine Maheo (Pitié-Salpêtrière Hospital, Paris, clinical research assistant), Manon Gomes (Pitié-Salpêtrière Hospital, Paris, clinical research assistant)

**Study sponsoring:** The ICEBERG Study was funded by the Programme d'investissements d’avenir (ANR-10-IAIHU-06), the Paris Institute of Neurosciences – IHU (IAIHU-06), the Agence Nationale de la Recherche (ANR-11-INBS-0006), and Électricité de France (Fondation d’Entreprise EDF).

#### NCER-PD/LuxPARK consortium

We would like to thank all participants of the Luxembourg Parkinson's Study for their important support to our research. Furthermore, we acknowledge the joint effort of the National Centre of Excellence in Research on Parkinson's Disease (NCER-PD) Consortium members from the partner institutions Luxembourg Centre for Systems Biomedicine, Luxembourg Institute of Health, Centre Hospitalier de Luxembourg, and Laboratoire National de Santé generally contributing to the Luxembourg Parkinson's Study as listed below:

Geeta ACHARYA[2], Gloria AGUAYO[2], Myriam ALEXANDRE[2], Muhammad ALI[1], Wim AMMERLANN[2], Giuseppe ARENA[1], Rudi BALLING[1], Michele BASSIS[1], Katy BEAUMONT[2], Regina BECKER[1], Camille BELLORA[2], Guy BERCHEM[3], Daniela BERG[11], Alexandre BISDORFF[5], Ibrahim BOUSSAAD[1], Kathrin BROCKMANN[11], Jessica CALMES[2], Lorieza CASTILLO[2], Gessica CONTESOTTO[2], Nico DIEDERICH[3], Rene DONDELINGER[5], Daniela ESTEVES[2], Guy FAGHERAZZI[2], Jean-Yves FERRAND[2], Manon GANTENBEIN[2], Thomas GASSER[11], Piotr GAWRON[1], Soumyabrata GHOSH[1], Marijus GIRAITIS[2],[3], Enrico GLAAB[1], Elisa GÓMEZ DE LOPE[1], Jérôme GRAAS[2], Mariella GRAZIANO[17], Valentin GROUES[1], Anne GRÜNEWALD[1], Wei GU[1], Gaël HAMMOT[2], Anne-Marie HANFF[2,20,21], Linda HANSEN[1,3], Michael HENEKA[1], Estelle HENRY[2], Sylvia HERBRINK[6], Sascha HERZINGER[1], Michael HEYMANN[2], Michele HU[8], Alexander HUNDT[2], Nadine JACOBY[18], Jacek JAROSLAW LEBIODA[1], Yohan JAROSZ[1], Sonja JÓNSDÓTTIR[2], Quentin KLOPFENSTEIN[1], Jochen KLUCKEN[1,2,3], Rejko KRÜGER[1,2,3], Pauline LAMBERT[2], Zied LANDOULSI[1], Roseline LENTZ[7], Inga LIEPELT[11], Robert LISZKA[14], Laura LONGHINO[3], Victoria LORENTZ[2], Paula Cristina LUPU[2], Tainá M. MARQUES[1], Clare MACKAY[10], Walter MAETZLER[15], Katrin MARCUS[13], Guilherme MARQUES[2], Patricia MARTINS CONDE[1], Patrick MAY[1], Deborah MCINTYRE[2], Chouaib MEDIOUNI[2], Francoise MEISCH[1], Myriam MENSTER[2], Maura MINELLI[2], Michel MITTELBRONN[1,4], Brit MOLLENHAUER[12], Friedrich MÜHLSCHLEGEL[4], Romain NATI[3], Ulf NEHRBASS[2], Sarah NICKELS[1], Beatrice NICOLAI[3], Jean-Paul NICOLAY[19], Fozia NOOR[2], Marek OSTASZEWSKI[1], Clarissa P. C. GOMES[1], Sinthuja PACHCHEK[1], Claire PAULY[1,3], Laure PAULY[2,20], Lukas PAVELKA[1,3], Magali PERQUIN[2], Nancy E. RAMIA[1,] Rosalina RAMOS LIMA[2], Armin RAUSCHENBERGER[1], Rajesh RAWAL[1], Dheeraj REDDY BOBBILI[1], Kirsten ROOMP[1], Eduardo ROSALES[2], Isabel ROSETY[1], Estelle SANDT[2], Stefano SAPIENZA[1], Venkata SATAGOPAM[1], Margaux SCHMITT[2], Sabine SCHMITZ[1], Reinhard SCHNEIDER[1], Jens SCHWAMBORN[1], Amir SHARIFY[2], Ekaterina SOBOLEVA[1], Kate SOKOLOWSKA[2], Hermann THIEN[2], Elodie THIRY[3], Rebecca TING JIIN LOO[1], Christophe TREFOIS[1], Johanna TROUET[2], Olena TSURKALENKO[2], Michel VAILLANT[2], Mesele VALENTI[2], Gilles VAN CUTSEM[1,3], Carlos VEGA[1], Liliana VILAS BOAS[3], Maharshi VYAS[1], Richard WADE-MARTINS[9], Paul WILMES[1], Evi WOLLSCHEID-LENGELING[1], Gelani ZELIMKHANOV[3]

1. Luxembourg Centre for Systems Biomedicine, University of Luxembourg, Esch-sur-Alzette, Luxembourg

2. Luxembourg Institute of Health, Strassen, Luxembourg
3. Centre Hospitalier de Luxembourg, Strassen, Luxembourg
4. Laboratoire National de Santé, Dudelange, Luxembourg
5. Centre Hospitalier Emile Mayrisch, Esch-sur-Alzette, Luxembourg
6. Centre Hospitalier du Nord, Ettelbrück, Luxembourg
7. Parkinson Luxembourg Association, Leudelange, Luxembourg
8. Oxford Parkinson's Disease Centre, Nuffield Department of Clinical Neurosciences, University of Oxford, Oxford, UK
9. Oxford Parkinson's Disease Centre, Department of Physiology, Anatomy and Genetics, University of Oxford, South Parks Road, Oxford, UK
10. Oxford Centre for Human Brain Activity, Wellcome Centre for Integrative Neuroimaging, Department of Psychiatry, University of Oxford, Oxford, UK
11. Center of Neurology and Hertie Institute for Clinical Brain Research, Department of Neurodegenerative Diseases, University Hospital Tübingen, Germany
12. Paracelsus-Elena-Klinik, Kassel, Germany
13. Ruhr-University of Bochum, Bochum, Germany
14. Westpfalz-Klinikum GmbH, Kaiserslautern, Germany
15. Department of Neurology, University Medical Center Schleswig-Holstein, Kiel, Germany
16. Department of Neurology Philipps, University Marburg, Marburg, Germany
17. Association of Physiotherapists in Parkinson's Disease Europe, Esch-sur-Alzette, Luxembourg
18. Private practice, Ettelbruck, Luxembourg
19. Private practice, Luxembourg, Luxembourg
20. Faculty of Science, Technology and Medicine, University of Luxembourg, Esch-sur-Alzette, Luxembourg
21. Department of Epidemiology, CAPHRI School for Public Health and Primary Care, Maastricht University Medical Centre+, Maastricht, the Netherlands
